# Lopinavir-ritonavir alone or combined with arbidol in the treatment of 73 hospitalized patients with COVID-19: a pilot retrospective study

**DOI:** 10.1101/2020.04.25.20079079

**Authors:** Xiu Lan, Chuxiao Shao, Xu Zeng, Zhenbo Wu, Yanyan Xu

## Abstract

**Objectives:** This study aimed to evaluate the antiviral efficacy of lopinavir/ritonavir alone or combined with arbidol in the treatment of hospitalized patients with common coronavirus disease-19 (COVID-19).

**Methods:** In this retrospective observational study, COVID-19 hospitalized patients were identified and divided into two groups based on the antiviral agents used during their hospitalization. Group-LR patients were treated with single antiviral drug of lopinavir-ritonavir. Group-LR+Ar patients were treated with lopinavir-ritonavir combined with arbidol for antiviral therapy at least 3 days. Patients were assessed for different clinical outcomes.

**Results:** A total of 34 and 39 patients were identified for Group-LR and Group-LR+Ar, respectively. Treatment with lopinavir–ritonavir alone was not difference from lopinavir-ritonavir combined with arbidol in overall cure rate of COVID-19 hospitalized patients (92.3% and 97.1%, respectively). In a modified intention-to-treat analysis, lopinavir–ritonavir combined with abidol led to a median time of hospital stay that was shorter by 1.5 days than group-LR (12.5 days vs. 14 days). The percentages of COVID-19 RNA clearance was 92.3 in group-LR and 97.1 in group-LR+Ar. The mean time of virus turning negative was 11.5±9.0 days in group-LR+Ar that were longer than group-LR. Treatment of lopinavir-ritonavir combined with arbidol did not significantly accelerate main symptoms improvement and promote the image absorption of pulmonary inflammation.

**Conclusion:** No benefit was observed in the anti-virus effect of lopinavir–ritonavir combined with arbidol compared with lopinavir-ritonavir alone in the hospitalized patients with COVID-19. More clinical observations in COVID-19 patients may help to confirm or exclude the effect of antiviral agents.

## 1. Introduction

COVID-19 is a new infectious disease. Since the COVID-19 outbreak, the number of confirmed cases of the virus reached 82052 on the Chinese mainland as of the mid-April, and 3339 cases died[1]. Among them, the cumulative confirmed cases in Hubei Province and Wuhan accounted for 74.3% and 43.2% of the total cases in China, respectively, which is the top priority of epidemic prevention and control[2]. Wang et al[3] reported 138 novel coronavirus pneumonia hospitalized patients’ clinical characteristics in Wuhan, China. Patients’ common symptoms included of fever, fatigue and dry cough. Lymphopenia, prolonged prothrombin time and elevated lactated hydrogenase were occurred in most of hospitalized patients with COVID-19. With the COVID-19 spread rapidly throughout China, Guan et al[4] extracted data from 1099 patients in 30 provinces of mainland China reported that novel coronavirus pneumonia patients often presented without fever, and many did not have abnormal radiologic findings. The understanding of novel coronavirus pneumonia is a gradual exploration process. As so for, prevention and control protocol for Novel Coronavirus Pneumonia issued by General Office of National Health Commission have been updated to the version 7.

There is still no specific medicine for novel coronavirus pneumonia. COVID-19 is categorized as beta genus coronavirus, same as the two other strains-human severe acute respiratory syndrome coronavirus (SARS-Cov) and Middle East respiratory syndrome coronavirus (MERS-Cov)[3]. Antiviral agents recommended in prevention and control protocol for Novel Coronavirus Pneumonia (version 7) was mainly summarized by the antiviral therapy of MERS-Cov and SARS-Cov [6]. Lopinavir-ritonavir is an HIV protease inhibitors and its registered therapeutics is for the treatment of HIV infection. Lopinavir is screened from FDA-approved compound library that can inhibit MERS-CoV replication in the low-micromolar range. Moreover, lopinavir can also inhibit the replication of SARS coronavirus and human coronavirus 229E[7]. Arbidol is a RNA synthesis inhibitor which is used for the prophylaxis and treatment of Influenza A and B, SARS, Hepatitis B and C, Herpies, Dengue Fever, and RSV. Abidol have been identified effective with anti-coronavirus activity in vitro[8]. Therefore, the efficacy and safety of antiviral drugs in COVID-19 need to be further confirmed by the clinical trials. Meanwhile, there is little information of clinical antiviral therapy on the lopinavir-ritonavir alone or combined with arbidol in the COVID-19 patients. In the present study, we retrospective studied medical records of 73 hospitalized patients with COVID-19 and evaluated the antivirus efficacy of lopinavir/ritonavir alone or combined with arbidol.

## 2. Methods

### 2.1. Study Design and Participants

The protocol was approved by the Ethics Committee of Lishui Hospital of Zhejiang University (Ethical Review of Clinical Research-2020-5) and was registered in Chinese Clinical Trail Registry (ChiCTR2000030391). Cases material of discharged patients were collected from February 21 to March 18, 2020 at Lishui Central Hospital, Zhejiang, China and Wuhan fourth hospital, Hubei, China, which is a designated hospital for COVID-19. All patients were diagnosed and treated according to the diagnosis and treatment protocol for novel coronavirus pneumonia issued by General Office of National Health Commission[5]. Inclusion criteria: (1) hospitalized patients with laboratory-confirmed COVID-19; (2) hospitalized patients with COVID-19 were treated with lopinavir-ritonavir alone or combined with arbidol for antiviral therapy. Exclusion criteria: (1) hospitalized patients with pneumonia caused by other pathogens; (2) hospitalized patients with COVID-19 haven’t received lopinavir-ritonavir for antiviral therapy.

A total of 73 cases were eligible to participate which meet the inclusion criteria and exclusion criteria. 34 of the 73 patients who had been treated with lopinavir-ritonavir (400 mg and 100mg, orally) twice daily were identified to the Group-LR, and the remainder who had been treated with lopinavir-ritonavir (400 mg and 100mg, orally) twice daily combined with arbidol (200 mg, orally) three times a day was identified to the Group-LR+Ar. All eligible patients received supportive treatment if necessary, such as supplemental oxygen, noninvasive and invasive ventilation, antibiotic agents and vasopressor support.

### 2.2. Data collection

Epidemiological, demographic, clinical, laboratory, radiological characteristics, treatment, and outcome data were collected through a review of electronic medical records. The data were reviewed by a trained team of physicians. Information recorded included demographic data, clinical presentation, medical history, laboratory values (ie, hematological parameters, biochemical parameters), time of virus negative, chest computed tomographic (CT) scans, and treatment measures during the hospital stay (ie, antiviral therapy, corticosteroid therapy, immunosuppressive therapy). The clinical classification of COVID-19 patients were carried out according to the diagnosis and treatment protocol for novel coronavirus pneumonia issued by General Office of National Health Commission[5]. The laboratory test results were grouped by admission time on the day 0-2, 3-5, 6-9, 10-14, and >15.

The primary end point was the rate of cure, defined as hospitalized patients with COVID-19 meet the discharge criteria. The discharge criteria were as follows[5]: 1) Body temperature is back to normal for more than three days; 2) Respiratory symptoms improve obviously; 3) Pulmonary imaging shows obvious absorption of inflammation; 4) Nuclei acid tests negative twice consecutively on respiratory tract samples such as sputum and nasopharyngeal swabs (sampling interval being at least 24 hours). Secondary end points were hospital stay, the rate of COVID-19 RNA clearance, the time of nucleic acid turning negative and symptoms disappeared to admission. The result and time SARS-COV-2 nucleic acid test of respiratory samples was subject to the last test. If the COVID-19 nucleic acid was negative for two consecutive tests, the first test time was used for nucleic acid turning negative.

### 2.3. Statistical Analysis

Categorical variables were described as frequency rates and percentages, and continuous variables were described using mean, median, and inter quartile range (IQR) values. Proportions for categorical variables were compared using the χ2 test, although the Fisher exact test was used when the data were limited. Continuous variables were expressed as means ± SD when the data were normally distributed, and means were compared using in dependent group t tests; otherwise, data (nonnormal distribution) were expressed as median and inter quartile range using the Mann-Whitney test. All statistical analyses were performed using SPSS (Statistical Package for Results the Social Sciences) version 16.0 software (SPSS Inc). *P* < 0.05 was considered to be significantly different between the two groups.

## 3. Results

### 3.1. Demographic and Clinical Characteristics

Of these 73 hospitalized patients with COVID-19 who was discharged from February 8 to March 13, 2020. The largest number of patients (60) had been admitted to Wuhan fourth hospital. The rest of patients were from Lishui Central Hospital. The demographic and clinical characteristics of the patients were shown in Table 1. The mean age of group-LR+Ar was 52.3±15.8 years (range, 21-81 years), and 26 (66.7%) were men. The mean age of group-LR was 59.5±13.6 years (range, 30-87 years), and 11 (32.4%) were men. The differences of mean age and sex ratio between group-LR+Ar and group-LR patients had statistically significant. Number of patients in all level of ages has no statistical difference. In the group-LR+Ar, the ratio of ordinary and heavy type of COVID-19 was 71.8% and 29.2%, respectively. The ratio of ordinary and heavy type of COVID-19 was 61.8% and 38.2% in the group-LR. Some patients had one or more coexisting medical conditions. Hypertension, coronary heart disease and diabetes were the most common comorbidities. 15(38.5%) patients had comorbidities in the group-LR+Ar, cardiovascular and cerebrovascular disease (8[20.5%]) and endocrine system disease (4[10.3%]). 19(55.9%) patients had comorbidities in the group-LR, cardiovascular and cerebrovascular disease (12[35.3%]) and endocrine system disease (6[17.6%]). The most common signs and symptoms were cough 27([69.2%]), fever (30[76.9%]), and chest tightness (10[25.6%]) in group-LR+Ar. The most common signs and symptoms were fever (28 [82.4%]), cough (25[73.5%]), and chest tightness (11[32.4%]) in group-LR+Ar. Less common symptoms were fatigue, diarrhea, headache, sore throat.

**Table 1.** Baseline characteristics of group-LR+Ar and group-LR

|  | No. (%) |  |  |
| --- | --- | --- | --- |
|  | LR+Ar (N=39) | LR (N=34) | P Value |
| Age, yr, mean ±SD | 52.3±15.8 | 59.5±13.6 | 0.043 |
| <65 yr | 29 | 20 | 0.159 |
| ≥65 yr | 10 | 14 | |
| Sex |  |  |  |
| Female | 13(33.3) | 23(67.6) | 0.003 |
| Male | 26(66.7) | 11(32.4) |  |
| Clinical classification |  |  |  |
| Ordinary | 28(71.8) | 21(61.8) | 0.363 |
| Heavy | 11(29.2) | 13(38.2) |  |
| Days from onset of symptoms to hospital admission, d, mean ±SD | 10.0±8.0 | 10.6±7.1 | 0.730 |
| Comorbidities | 15(38.5) | 19(55.9) | 0.137 |
| Any comorbidity | 24(61.5) | 15(44.1) |  |
| Cardiovascular and cerebrovascular disease | 8(20.5) | 12(35.3) | 0.158 |
| Endocrine system disease | 4(10.3) | 6(17.6) | 0.499 |
| Malignant tumor | 2(5.1) | 2(5.9) | >.99 |
| Respiratory system disease | 0 | 1(2.9) | 0.466 |
| Digestive system disease | 0 | 1(2.9) | 0.466 |
| Renal disease | 1(2.6) | 0 | >.99 |
| Liver disease | 1(2.6) | 0 | >.99 |
| Signs and symptoms |  |  |  |
| Fever | 27(69.2) | 28(82.4) | 0.194 |
| Cough | 30(76.9) | 25(73.5) | 0.737 |
| Chest tightness | 10(25.6) | 11(32.4) | 0.608 |
| Fatigue | 3(7.9) | 7(20.6) | 0.172 |
| Diarrhea | 3(7.9) | 5(14.7) | 0.460 |
| Others | 7(1.8) | 3(8.8) | 0.321 |

### 3.2. Primary outcome

The antiviral efficacy of lopinavir-ritonavir alone or combined with arbidol in the treatment of 73 hospitalized patients with COVID-19 were shown in Table 2. In the group-LR+Ar, 36(92.3%) patients were cured and discharged from hospital, and 2(5.1%) patients were admitted and transferred to the ICU because of the condition aggravation. In the group-LR, 33(97.1%) were cured and discharged from hospital with no patients were transferred to the ICU. The mortality was 2.6% and 2.9% in group-LR+Ar and group-LR, respectively. The rate of COVID-19 RNA clearance was 92.3% of group-LR+Ar and 97.1% of group-LR. Removed 3 patients who transferred to ICU or died in the group-LR+Ar, the time of nucleic acid turning negative was 11.5±9.0 days and hospital stay was 14.4±7.9 days. Removed 1 died case in the group-LR, the time of nucleic acid turning negative was 9.9±7.5 days and hospital stay was 16.0±9.0 days. Of the 39 patients in the group-LR+Ar, the pulmonary image manifestations of 33(84.6%) patients were absorbed at discharged, included of less absorption and marked improvement. 13(38.2%) patients were absorbed at discharged in the group-LR. No absorption of pulmonary image manifestations had 2 cases in each group. 4(10.3%) patients’ chest CT findings were worsened in group-LR+Ar and 1 case was worsened in group-LR. Fever, cough and chest tightnessd were the main symptoms in hospitalized patients with COVID-19. The disappearance time of fever, cough and chest tightnessd in the group-LR+Ar were 3.4±2.7, 9.0±6.2 and 6.0±2.1 days while they were 2.6±3.4, 6.6±5.0 and 10.1±3.0 days in the group-LR, respectively.

**Table 2.** Comparison of the primary end point and secondary end points between group-LR+Ar and group-LR

| Variable | LR+Ar (N=39) | LR (N=34) | P value |
| --- | --- | --- | --- |
| Clinical outcome |  |  |  |
| Cured | 36(92.3) | 33(97.1) | 0.618 |
| Aggravated | 2(5.1) | 0 | 0.495 |
| Died | 1(2.6) | 1(2.9) | >.99 |
| Hospital stay, days<sup>a</sup> | 14.4±7.9 | 16.0±9.0 | 0.431 |
| COVID-19 RNA clearance | 36(92.3) | 33(97.1) | 0.618 |
| The time of nucleic acid turning negative<sup>a</sup> | 11.5±9.0 | 9.9±7.5 | 0.585 |
| Chest CT findings at discharge |  |  |  |
| Absorption | 33(84.6) | 31(91.1) | 0.489 |
| No change | 2(5.1) | 2(5.9) | >.99 |
| Advances | 4(10.3) | 1(2.9) | 0.363 |
| Main symptoms |  |  |  |
| Fever<sup>b</sup> | 3.4±2.7 | 2.6±3.4 | 0.369 |
| Cough<sup>c</sup> | 9.0±6.2 | 6.6±5.0 | 0.122 |
| Chest tightness<sup>d</sup> | 6.0±2.1 | 10.1±3.0 | 0.689 |

<sup>a</sup> The time of nucleic acid turning negative and hospital length of stay was calculated for 36 patients in the LR+Ar group and 33 patients in the LR group.
<sup>b</sup> Fever was calculated for 25 patients in the LR+Ar group and 27 patients in the LR group.
<sup>c</sup> Cough was calculated for 27 patients in the LR+Ar group and 24 patients in the LR group.
<sup>d</sup> Chest tightness was calculated for 8 patients in the LR+Ar group and 11 patients in the LR group.

### 3.3. Laboratory Parameters

6 clinical laboratory parameters included of white blood cell, neutrophil, lymphocyte, monocyte, C-reactive protein and procalcitonin were tracked during their hospitalization. Dynamic changes of the laboratory parameters was classified on the day 1-2, 3-5, 6-9, 10-14 and after 15 since the patients admitted to hospital (Figure 1). There were no significant differences in 6 clinical laboratory parameters between group-LR+Ar and group-LR. Mean white blood cell and neutrophil count were in normal range in both two groups. The level of lymphocyte count decreased in both two groups that next to the lower normal limit, the minimum was at 3-5 days after admitted to hospital. Both of the two groups were in low level of lymphocyte at 0-15 days after hospitalization with no significant difference. On the contrary, the level of monocyte was next to the upper normal limit. The level of the inflammatory biomarker C-reactive protein was marked increased in early hospitalization of both two groups, and then gradually declined over time during hospitalization. Mean C-reactive protein of was higher in group-LR than that in group-LR+Ar. Mean value of procalcitonin was slightly higher than the upper normal limit with no significant difference between two groups.

**FIGURE 1.**
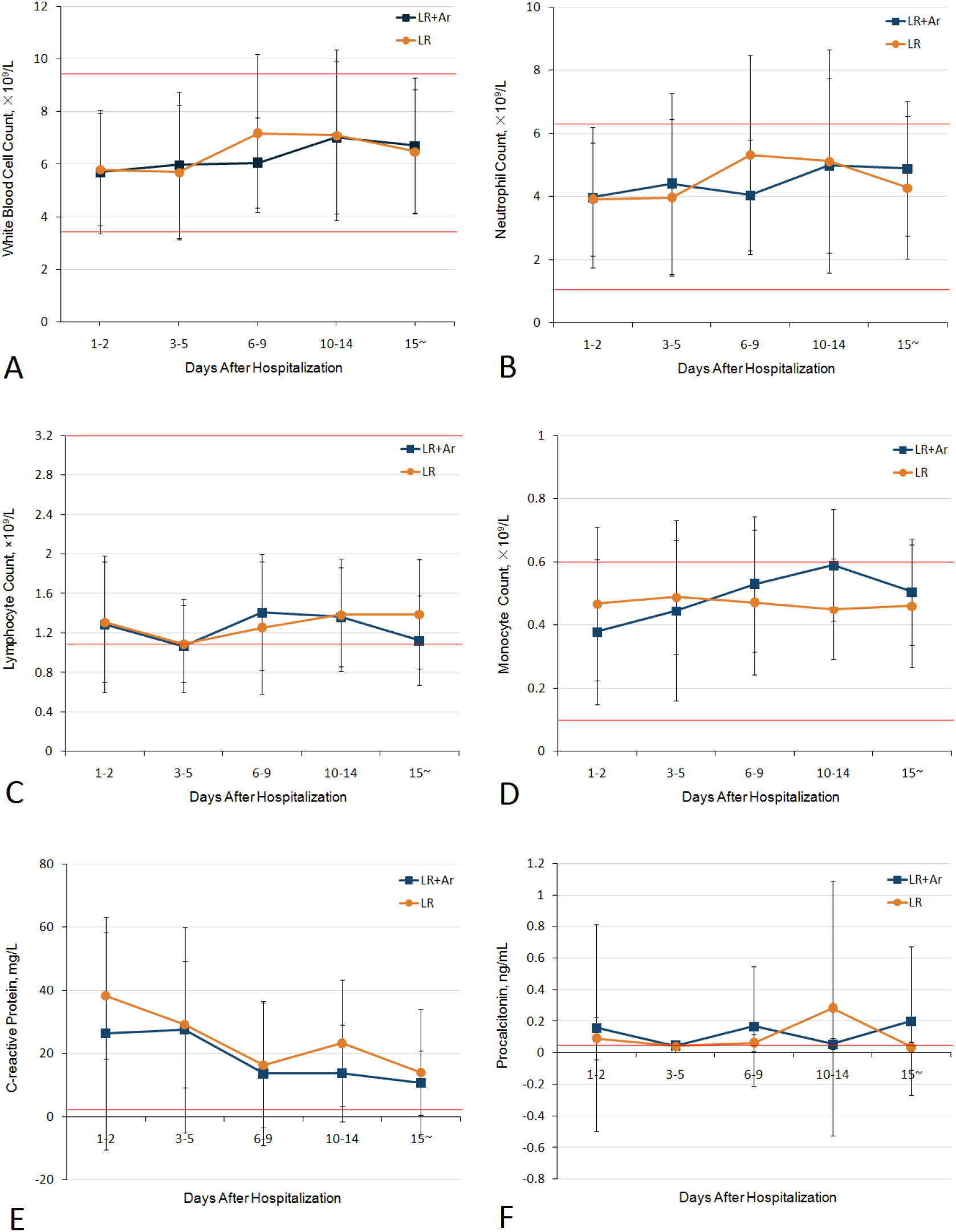
Dynamic profile of laboratory parameters in group-LR+Ar and group-LR after the patients admitted to hospital. (A) White blood cell count, normal range 3.5-9.5×10^9^/L. (B) Neutrophil count, normal range 1.8-6.3×10^9^/L. (C) Lymphocyte count, normal range 1.1-3.2×10^9^/L. (D) Monocyte count, normal range 0.1-0.6×10^9^/L. (E) C-reactive protein, normal range 0-3 mg/L. (F) Procalcitonin, normal range 0-0.05ng/mL.

## 4. Discussion

In this study, we found that there was no significant difference in the end points of COVID-19 patients in antiviral therapy, such as cure rate, hospitalization time, virus turning negative rate and the time of virus turning negative between the single antiviral drug of lopinavir-ritonavir and lopinavir-ritonavir combined with abidol. However, in the modified intention to-treat analysis, which excluded patients with early death and transferred to the ICU, the median hospitalization time of lopinavir-ritonavir combined with abidol group was shorten than lopinavir-ritonavir group (1.25 days vs. 14 days). Of note, the overall mortality of lopinavir-ritonavir combined with abidol group and lopinavir-ritonavir group were 2.6% and 2.9% in this study. It was significantly lower than the 11% to 22.1% mortality reported in previous studies of hospitalized patients with COVID-19[9-11]. The main reason was that there were fewer severe cases enrolled in this study. The ratio of heavy COVID-19 in lopinavir-ritonavir combined with abidol group and lopinavir-ritonavir group were 29.2% and 38.2%, resecptively.

According to the results, we did not find that two antiviral drugs lopinavir–ritonavir combined with abidol treatment improved clinical symptoms and accelerated virological inhibition compared with single antiviral drug lopinavir–ritonavir treatment. In contrast, the time virus turning negative and the duration time of fever and cough in lopinavir–ritonavir combined with abidol group were longer than lopinavir–ritonavir treatment group. Two consecutive nucleic acid tests were negative, and clinical symptom improvement is the discharge standard of COVID-19 patients. However, the clinical symptoms of some COVID-19 patients had improved,the results of multiple nucleic acid tests were positive and the longest viral shedding was 36 days. It was consistence with the previous study[12]. A recent report showed that the results of nucleic acid re-test of convalescent patients with COVID-19 were positive. Lan et al[13] found that the results of pharyngeal swab nucleic acid test were positive in the follow-up of 4 cured COVID-19 medical staff. Chen et al [14] reported that the proportion of turning positive was 3 / 7 and the interval of virus nucleic acid turning positive was 5-7 days in the follow-up study of 7 COVID-19 patients who met the national discharge standard.

COVID-19 is an acute or chronic pneumonia caused by SARS-COV-2 infection[15]. Pulmonary pathology of early phase SARS-COV-2 pneumonia exhibited exudative and proliferative phase acute lung injury such as edema, inflammatory infiltrate, pneumocyte hyperplasia, and organization[15]. Other studies found that the neutrophil count, D-dimer, blood urea, and creatinine levels continued to increase, and the lymphocyte counts continued to decrease in COVID-19 nonsurvivors[3]. Neutrophilia may be related to cytokine storm induced by virus invasion. We found that the level of leukocytes and neutrophils in patients with COVID-19 was normal, and the lymphocyte count was continued in lower level. This results were consistent with the previous report that COVID-19 accompany with lymphocytopenia[3,16]. COVID-19 may damage lymphocytes, especially T lymphocytes, and then damage the immune system during disease[16]. CRP is a useful gauge of inflammation. From the trend of CRP,CRP values of hospitalized patients with COVID-19 were increased slightly at admission and then decreased gradually. It is suggested that host defense against invading pathogens as well as inflammatory response. PCT is sensitive indicator of bacterial infection[17]. The profile of PCT levels was relatively stable with no markedly raise.

Our trial has several limitations. First, this study was a retrospective analysis with no randomized, future prospective study can overcome this defect. Second, there were only 73 COVID-19 cases in this study. According to the study data of lopinavir-ritonavir in the treatment of severe COVID-19 cases[11], we concluded that the sample size need 300 subjects to evaluate the efficacy of lopinavir-ritonavir combined with abidor was better or worse than that of the single drug treatment of lopinavir-ritonavir. Third, the age and gender of two groups were not balanced because of the small sample. However, there was no significant difference between groups after stratification of gender and age. In the future, increasing the number of research samples can improve the test efficiency and enhance the credibility of research results. Finally, the results of this study suggested that the overall treatment effect of COVID-19 was well. It was difficult to judge the antiviral effect using cure rate and virus turning negative rate as the primary endpoints. Therefore, it may be more feasible in clinical practice to find a more suitable COVID-19 population or primary endpoints for evaluating the therapeutic effect of antiviral drugs, such as evaluating whether it can reduce mortality in severely ill population.

In conclusion, we found that lopinavir–ritonavir alone or combined with arbidol treatment did not significantly increase cure rate, reduce hospitalization time, raise virus negative rate, and diminish virus negative time in hospitalized patients with COVID-19. These preliminary results could inform future trials of COVID-19 to evaluate this and other medication in the treatment of infection with SARS-CoV-2. Whether lopinavir–ritonavir combined with other antiviral drugs might enhance antiviral effects and improve clinical outcomes need more clinical observations.

## Data Availability

The data used to support the findings of this study have been deposited in the figshare repository (DOI:10.6084/m9.figshare.12195735).

https://figshare.com/articles/COVID19_inpatient_cases_data_xls/12195735

## Data Availability

The data used to support the findings of this study are available from the corresponding author upon request.

## Acknowledgements

This work was supported by the High Level Talented Person Cultivating Program of Lishui Science and Technology Bureau under Grant No.2018RC06, Zhejiang Provincial Natural Science Foundation of China under Grant no. LY18H300007, and Zhejiang TCM science and technology program under Grant No.2013ZB148.

